# Risk-Factors of Crimean-Congo Haemorrhagic Fever Across Six Tertiary Hospitals in Afghanistan from 2022 to 2025: Reported Exposures, Healthcare-Worker Clustering, and Ribavirin Treatment-Timing Analyses

**DOI:** 10.64898/2026.10.24.26366389

**Authors:** Rehmatullah Zadran, Mohammad Nadir Sahak, Fatima Arifi, Yiyu Chen, Husna Sadat, Ghayasuddin Saraj, Sanah Essayagh, Touria Essayagh, Lisa J. White, Nathaniel Hupert, Siyu Chen

**Affiliations:** National Infectious Disease Hospital Laboratory, Kabul, Afghanistan; Independent Researcher, Kabul, Afghanistan; Independent Researcher. Toronto, Canada; School of Biological and Behavioural Sciences, Queen Mary University of London, London, UK; National Disease Surveillance & Response, Kabul, Afghanistan; Hassan First University of Settat, Faculté des Sciences et Techniques, Laboratoire Agroalimentaire et Santé, Settat, Morocco; Hassan First University of Settat, Institut Supérieur des Sciences de la Santé, Laboratoire des Sciences et ingénierie biomédicales, biophysique et santé, Settat, Morocco; Model Health Ltd, Brixham, UK; Departments of Population Health Sciences and of Medicine, Weill Cornell Medicine, New York City, USA; Department of Health Data Science, University of Liverpool, UK.

## Abstract

**Introduction:** Crimean-Congo haemorrhagic fever (CCHF) is endemic in Afghanistan, but multi-hospital data characterising reported exposure patterns, healthcare-worker clustering, and ribavirin-mortality associations after accounting for treatment timing are lacking.

**Methods:** A retrospective cohort study included 737 laboratory-confirmed CCHF patients admitted to six tertiary hospitals in Afghanistan between January 2022 and December 2025. Reported exposures were assessed by occupation and tick-contact history. Clustering of healthcare-worker (HCW) infections during high caseload was assessed by permutation testing. Ribavirin-mortality association was assessed using multivariable Cox regression adjusted for clinical severity, age, sex, hospital, and comorbidity, with sensitivity analyses addressing treatment timing immortal-time bias.

**Results:** Among 737 patients, the median age was 35 years and 78.3% were male. The hospitalization fatality ratio (HFR) was 27.7% (204/737). Of 574 patients with recorded occupation: 33.6% were livestock handlers, 12.9% slaughterhouse workers or butchers, 24.9% household meat preparers, and only 11.1% reported tick exposure. Nineteen patients were HCWs, and 18 became infected during periods of concurrent CCHF admissions, and clustering exceeded chance (p=0.012), consistent with healthcare-associated exposure during surges. Personal protective equipment (PPE) was used in 728 (98.6%) admissions, but its composition was incomplete: 532 (72.2%) comprised mask and gloves without a gown, and only 205 (27.8%) with mask, gloves, and gown. Isolation rooms were available for only 274 of 737 admissions (37.2%). Among 727 patients in the ribavirin analysis, 85.0% received treatment. Conventional analysis suggested lower mortality (adjusted HR 0.57) but affected by immortal-time bias. The association weakened for same-day treatment versus no treatment (HR 0.63, 95% CI 0.41–0.96; sensitive to model specification) and was absent in time-varying analysis (HR 0.93).

**Conclusion:** CCHF in this hospitalised Afghan cohort was commonly associated with reported animal and livestock-related exposure, while healthcare-worker infections clustered during outbreak surges. Isolation capacity and caseload may be important correlates of healthcare-associated risk alongside PPE availability. Ribavirin findings were sensitive to treatment-timing adjustment and cannot establish a causal survival benefit.

**Key Questions:**

- What is already known on this topic
  - CCHF is endemic in Afghanistan, but previous studies lacked individual-level multi-hospital data on reported transmission exposures, formal assessment of healthcare-worker clustering, and ribavirin treatment-timing analyses, while evidence from other endemic countries has been inconsistent.
- What this study adds
  - In 737 laboratory-confirmed cases across six tertiary hospitals, reported animal and livestock-related exposure was far more common than reported tick exposure, healthcare-worker infections clustered significantly during periods of high CCHF caseload and where isolation capacity was limited, and the apparent mortality benefit of ribavirin was substantially attenuated after accounting for treatment timing and immortal-time bias.
- How this study might affect research, practice or policy
  - The findings support stronger public health interventions such as One Health prevention at the animal-human interface, investment in dedicated isolation capacity together with complete PPE during outbreak surges, protective equipment during slaughter and butchering, veterinary surveillance of viraemic livestock, and risk-reduction before Eid-al-Adha.

## Introduction

Crimean-Congo haemorrhagic fever (CCHF), caused by an orthonairovirus transmitted primarily by *Hyalomma* ticks, is one of the most widely distributed viral haemorrhagic fevers, with documented human infections across more than 30 countries in Africa, the Balkans, the Middle East, and Central Asia [1–5]. Case fatality rates in hospitalised patients range from 5% to 40%, varying by clinical setting, severity distribution, and access to supportive care [6–7]. The global burden of CCHF is substantially underestimated due to surveillance gaps in endemic low-income countries, where diagnostic capacity and reporting infrastructure are limited [3, 8–9].

The geographic range of CCHF is expanding due to climate change, rising temperatures, land-use change, and migratory bird dispersal of the *Hyalomma* tick vectors, which have driven range expansion into previously non-endemic regions, including autochthonous transmission in Spain since 2016 [10] and the detection of established *Hyalomma marginatum* populations across southern Europe [11–12]. Climate projections indicate continued northward spread under moderate warming scenarios [13–14]. In parallel, endemic regions face escalating case burdens: Afghanistan [15–16], Türkiye [17–18], Iran (Islamic Republic of) [19] and Pakistan [20], have all reported increasing incidence over the past decade, attributed to growing unregulated movement and unsafe handling of livestock populations, disrupted vector control programmes, and fragile health systems during periods of conflict and political instability.

Afghanistan has reported an increasing burden of CCHF in the past decade. CCHF has been endemic since at least 1998, with spring-summer epidemic cycles driven by overlapping livestock handling and tick activity seasons [15–16]. National surveillance data from Afghanistan report crude case fatality rates ranging from approximately 8% to over 40%, depending on period and case definition with marked heterogeneity across provinces and documented nosocomial outbreaks [16, 21–23]. The 2025 season has produced the largest recorded CCHF outbreak in Afghanistan to date, with 1,407 suspected cases and 97 deaths reported by late October (CFR 6.9%), substantially exceeding the 2022–2024 three-year average and compounding pressure on a fragile healthcare system serving a population with limited access to diagnostics, intensive care, and uninterrupted drug supply chains [22].

Three interrelated clinical and epidemiological questions remain inadequately characterised in the Afghanistan context. First, although *Hyalomma* tick bites are classically implicated in CCHFV transmission, the relative contribution of direct contact with infected animal blood and tissues through occupational slaughter, butchering, and domestic meat preparation has not been documented. In resource-limited settings where tick surveillance is minimal to absent, understanding reported exposure pattern is essential for designing prevention programmes while acknowledging that tick bites may be unnoticed or unrecalled. Second, healthcare-associated CCHF infection is well-documented in other countries in the region, but formal evidence on outbreak-period clustering and associated health-system constraints in Afghanistan hospital settings remains limited [23–25]. This is the first multi-centre hospital study including data from all CCHF-treating hospitals in Afghanistan. Third, ribavirin is widely recommended for CCHF treatment by WHO, but observational evidence remains mixed and is vulnerable to confounding, facility-level differences, and immortal-time bias. Severity-stratified ribavirin-mortality associations from routine clinical data in Afghanistan have not previously been reported [26–28].

The objectives of this study are to characterize reported CCHF exposure patterns, healthcare-worker infection clustering, and ribavirin-mortality associations in a multi-centre hospital setting.

## Methods

### Study design and setting

We conducted a retrospective analysis of multi-hospital surveillance and hospital medical records data of all admitted patients with documented CCHF from six public tertiary hospitals in Afghanistan: Balkh Regional Hospital (n=101), Herat Infectious Disease Hospital (n=103), Kunduz Regional Hospital (n=47), Kandahar Regional Hospital (n=39), Nangarhar Regional Hospital (n=52), and the National Infectious Disease Hospital, Kabul (n=395). These hospitals are the regional designated centres for high threat pathogens in Afghanistan. Patients admitted between January 2022 and December 2025, were included. Diagnosis was established by RT-PCR, antigen detection, or ELISA IgM/IgG serology, following the WHO cascade diagnostic pathway. Ethics approval was obtained from Institutional Review Board of Afghanistan National Public Health Institute [A-3-475]; all data were de-identified prior to analysis (see case report form in Supplementary File 1).

### Participants

All 737 hospitalised patients with laboratory-confirmed CCHF were included. The study was a census of all eligible cases during the study period; no formal sample-size calculation was performed. Demographic information, occupation, residential setting, tick exposure history, comorbidities, clinical features, supportive treatments received, and final outcome (survival or in-hospital death) were abstracted from standardised case record forms. Occupation was recorded for 574 of 737 patients (77·9%); free-text entries were mapped to five pre-specified categories (farmer or livestock handler; healthcare worker; housewife or household caregiver including meat handling; slaughterhouse worker or butcher; jobless or other). Clinical severity was classified by attending physicians according to clinical criteria as mild, moderate, or severe based on clinical symptoms, haemorrhagic manifestations and laboratory parameters at admission. The severity grading score was used as the method for severity classification as based on earlier studies in Turkey [29].

### Statistical Analysis

All analyses were conducted in R Statistical Software (Version 4.3; R Core Team, 2026). Survival analyses used the survival package. Receiver operating characteristic analyses used the pROC package. Spatial analyses used the sf and ggplot2 packages. Continuous variables are reported as median (interquartile range) and categorical variables as count (percentage). Denominators are stated for all proportions and are restricted to patients with the variable recorded; the number of missing observations for each variable is given in the footnote to Table 1. Date fields were recorded in two encodings within the source database and were parsed with explicit day-first interpretation before any interval was derived.

**Table 1.**
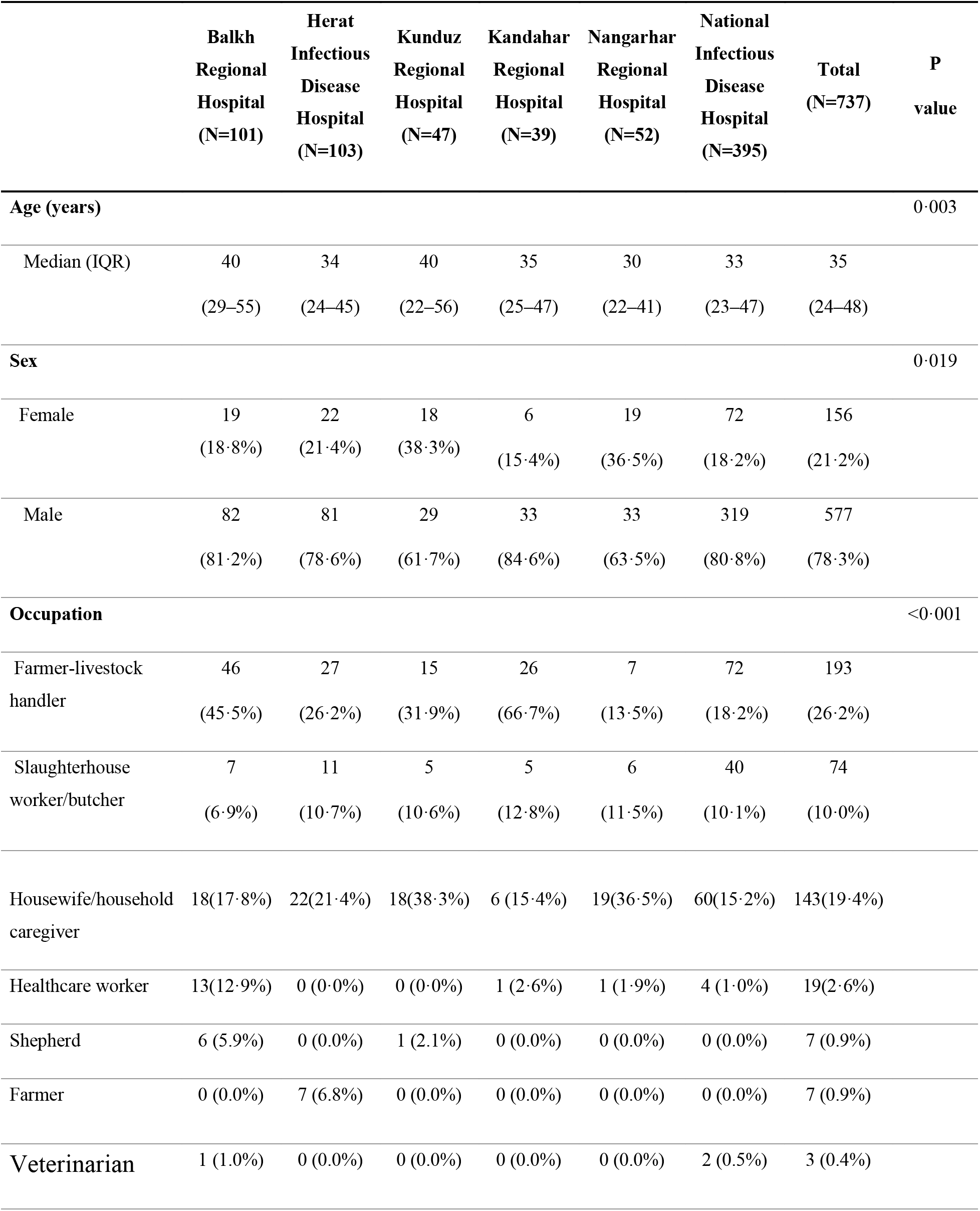
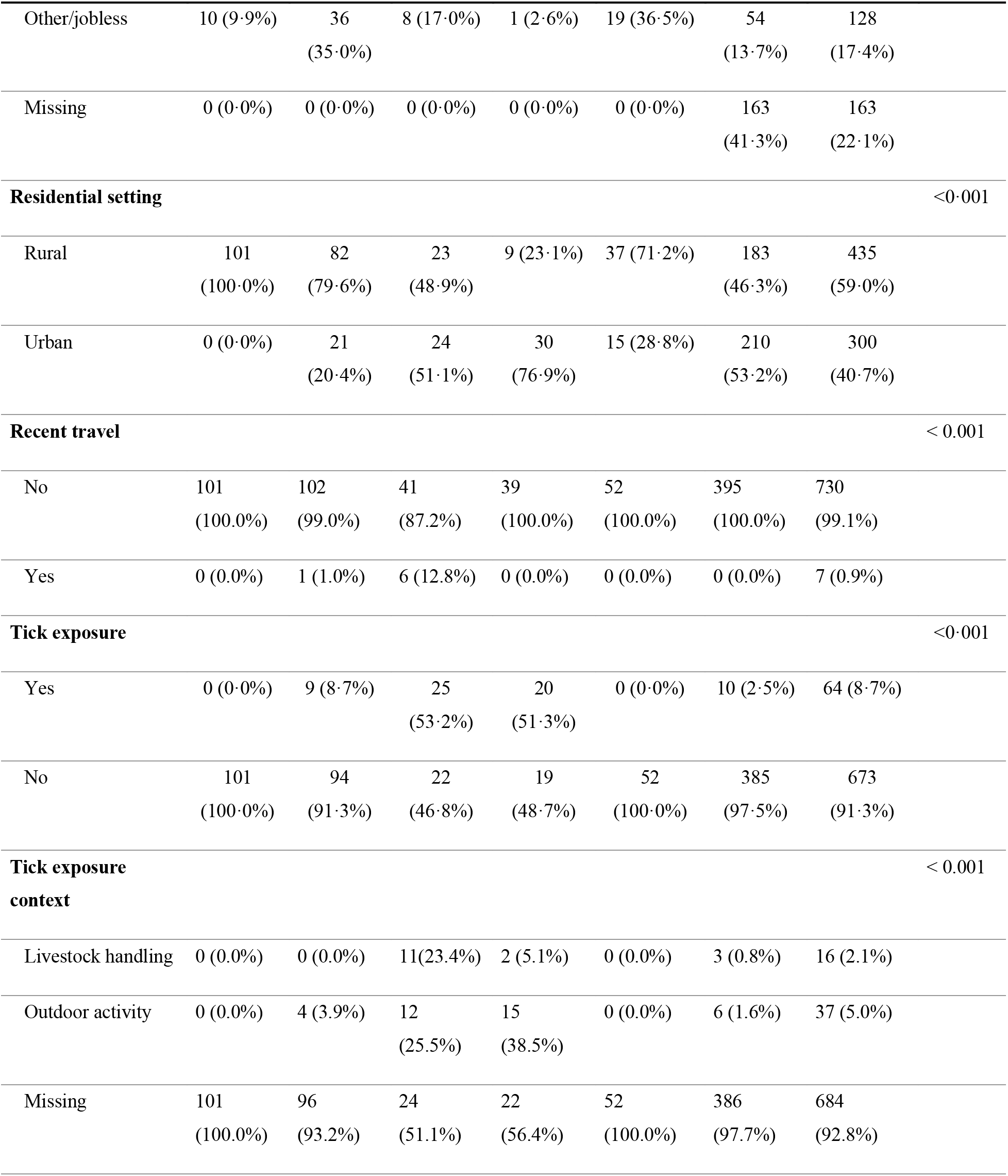
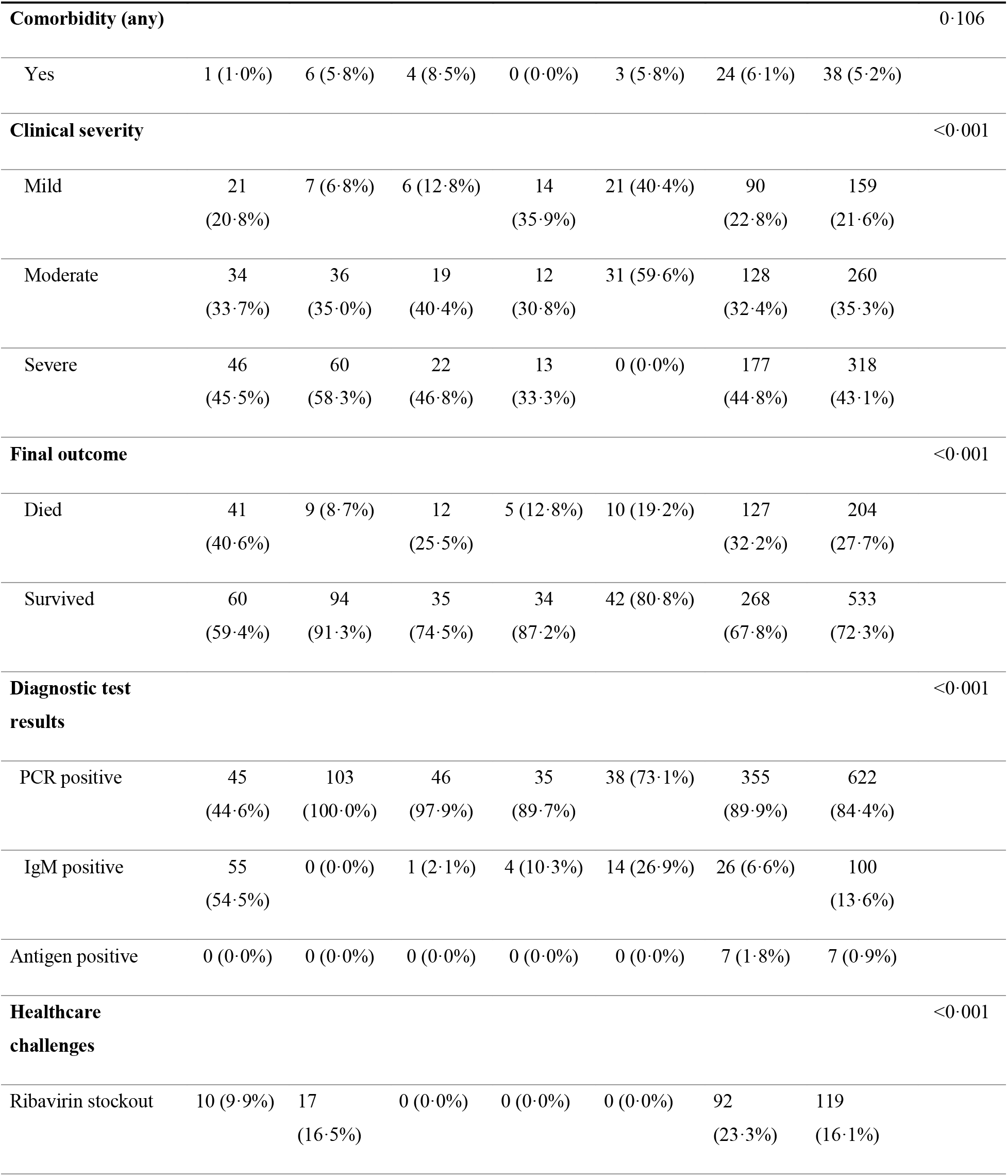
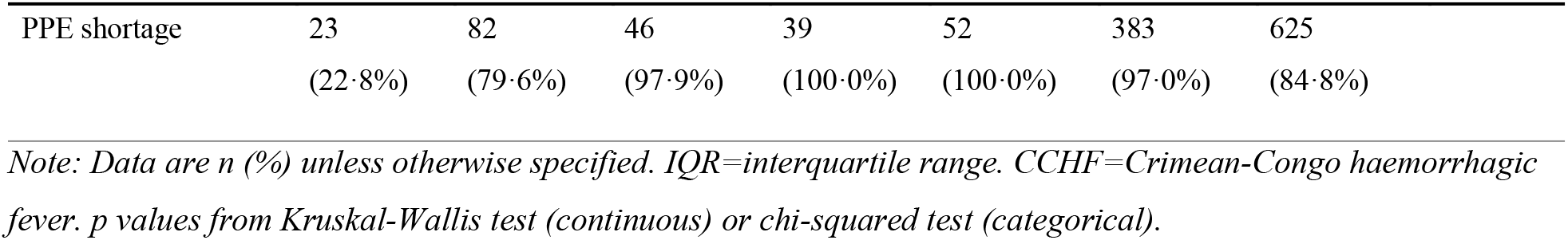
Demographic and epidemiological characteristics of 737 hospitalized patients with confirmed CCHF in Afghanistan, 2022-2025.

|  | **Balkh**<br>**Regional**<br>**Hospital**<br>**(N=101)** | **Herat**<br>**Infectious**<br>**Disease**<br>**Hospital**<br>**(N=103)** | **Kunduz**<br>**Regional**<br>**Hospital**<br>**(N=47)** | **Kandahar**<br>**Regional**<br>**Hospital**<br>**(N=39)** | **Nangarhar**<br>**Regional**<br>**Hospital**<br>**(N=52)** | **National**<br>**Infectious**<br>**Disease**<br>**Hospital**<br>**(N=395)** | **Total**<br>**(N=737)** | **P**<br>**value** |
| --- | --- | --- | --- | --- | --- | --- | --- | --- |
| **Age (years)** |  |  |  |  |  |  |  | 0.003 |
| Median (IQR) | 40<br>(29-55) | 34<br>(24-45) | 40<br>(22-56) | 35<br>(25-47) | 30<br>(22-41) | 33<br>(23-47) | 35<br>(24-48) |  |
| **Sex** |  |  |  |  |  |  |  | 0.019 |
| Female | 19<br>(18.8%) | 22<br>(21.4%) | 18<br>(38.3%) | 6<br>(15.4%) | 19<br>(36.5%) | 72<br>(18.2%) | 156<br>(21.2%) |  |
| Male | 82<br>(81.2%) | 81<br>(78.6%) | 29<br>(61.7%) | 33<br>(84.6%) | 33<br>(63.5%) | 319<br>(80.8%) | 577<br>(78.3%) |  |
| **Occupation** |  |  |  |  |  |  |  | <0.001 |
| Farmer-livestock<br>handler | 46<br>(45.5%) | 27<br>(26.2%) | 15<br>(31.9%) | 26<br>(66.7%) | 7<br>(13.5%) | 72<br>(18.2%) | 193<br>(26.2%) |  |
| Slaughterhouse<br>worker/butcher | 7<br>(6.9%) | 11<br>(10.7%) | 5<br>(10.6%) | 5<br>(12.8%) | 6<br>(11.5%) | 40<br>(10.1%) | 74<br>(10.0%) |  |
| Housewife/household<br>caregiver | 18(17.8%) | 22(21.4%) | 18(38.3%) | 6 (15.4%) | 19(36.5%) | 60(15.2%) | 143(19.4%) |  |
| Healthcare worker | 13(12.9%) | 0 (0.0%) | 0 (0.0%) | 1 (2.6%) | 1 (1.9%) | 4 (1.0%) | 19(2.6%) |  |
| Shepherd | 6 (5.9%) | 0 (0.0%) | 1 (2.1%) | 0 (0.0%) | 0 (0.0%) | 0 (0.0%) | 7 (0.9%) |  |
| Farmer | 0 (0.0%) | 7 (6.8%) | 0 (0.0%) | 0 (0.0%) | 0 (0.0%) | 0 (0.0%) | 7 (0.9%) |  |
| Veterinarian | 1 (1.0%) | 0 (0.0%) | 0 (0.0%) | 0 (0.0%) | 0 (0.0%) | 2 (0.5%) | 3 (0.4%) |  |
| Other/jobless | 10 (9.9%) | 36<br>(35.0%) | 8 (17.0%) | 1 (2.6%) | 19 (36.5%) | 54<br>(13.7%) | 128<br>(17.4%) |  |
| Missing | 0 (0.0%) | 0 (0.0%) | 0 (0.0%) | 0 (0.0%) | 0 (0.0%) | 163<br>(41.3%) | 163<br>(22.1%) |  |
| **Residential setting** |  |  |  |  |  |  |  | <0.001 |
| Rural | 101<br>(100.0%) | 82<br>(79.6%) | 23<br>(48.9%) | 9 (23.1%) | 37 (71.2%) | 183<br>(46.3%) | 435<br>(59.0%) |  |
| Urban | 0 (0.0%) | 21<br>(20.4%) | 24<br>(51.1%) | 30<br>(76.9%) | 15 (28.8%) | 210<br>(53.2%) | 300<br>(40.7%) |  |
| **Recent travel** |  |  |  |  |  |  |  | < 0.001 |
| No | 101<br>(100.0%) | 102<br>(99.0%) | 41<br>(87.2%) | 39<br>(100.0%) | 52<br>(100.0%) | 395<br>(100.0%) | 730<br>(99.1%) |  |
| Yes | 0 (0.0%) | 1 (1.0%) | 6 (12.8%) | 0 (0.0%) | 0 (0.0%) | 0 (0.0%) | 7 (0.9%) |  |
| **Tick exposure** |  |  |  |  |  |  |  | <0.001 |
| Yes | 0 (0.0%) | 9 (8.7%) | 25<br>(53.2%) | 20<br>(51.3%) | 0 (0.0%) | 10 (2.5%) | 64 (8.7%) |  |
| No | 101<br>(100.0%) | 94<br>(91.3%) | 22<br>(46.8%) | 19<br>(48.7%) | 52<br>(100.0%) | 385<br>(97.5%) | 673<br>(91.3%) |  |
| **Tick exposure**<br>**context** |  |  |  |  |  |  |  | < 0.001 |
| Livestock handling | 0 (0.0%) | 0 (0.0%) | 11(23.4%) | 2 (5.1%) | 0 (0.0%) | 3 (0.8%) | 16 (2.1%) |  |
| Outdoor activity | 0 (0.0%) | 4 (3.9%) | 12<br>(25.5%) | 15<br>(38.5%) | 0 (0.0%) | 6 (1.6%) | 37 (5.0%) |  |
| Missing | 101<br>(100.0%) | 96<br>(93.2%) | 24<br>(51.1%) | 22<br>(56.4%) | 52<br>(100.0%) | 386<br>(97.7%) | 684<br>(92.8%) |  |

|  | Balkh<br>Regional<br>Hospital<br>(N=101) | Herat<br>Infectious<br>Disease<br>Hospital<br>(N=103) | Kunduz<br>Regional<br>Hospital<br>(N=47) | Kandahar<br>Regional<br>Hospital<br>(N=39) | Nangarhar<br>Regional<br>Hospital<br>(N=52) | National<br>Infectious<br>Disease<br>Hospital<br>(N=395) | Total<br>(N=737) | P<br>value |
| --- | --- | --- | --- | --- | --- | --- | --- | --- |
| **Comorbidity (any)** |  |  |  |  |  |  |  | 0.106 |
| Yes | 1 (1.0%) | 6 (5.8%) | 4 (8.5%) | 0 (0.0%) | 3 (5.8%) | 24 (6.1%) | 38 (5.2%) |  |
| **Clinical severity** |  |  |  |  |  |  |  | <0.001 |
| Mild | 21<br>(20.8%) | 7 (6.8%) | 6 (12.8%) | 14<br>(35.9%) | 21 (40.4%) | 90<br>(22.8%) | 159<br>(21.6%) |  |
| Moderate | 34<br>(33.7%) | 36<br>(35.0%) | 19<br>(40.4%) | 12<br>(30.8%) | 31 (59.6%) | 128<br>(32.4%) | 260<br>(35.3%) |  |
| Severe | 46<br>(45.5%) | 60<br>(58.3%) | 22<br>(46.8%) | 13<br>(33.3%) | 0 (0.0%) | 177<br>(44.8%) | 318<br>(43.1%) |  |
| **Final outcome** |  |  |  |  |  |  |  | <0.001 |
| Died | 41<br>(40.6%) | 9 (8.7%) | 12<br>(25.5%) | 5 (12.8%) | 10 (19.2%) | 127<br>(32.2%) | 204<br>(27.7%) |  |
| Survived | 60<br>(59.4%) | 94<br>(91.3%) | 35<br>(74.5%) | 34<br>(87.2%) | 42 (80.8%) | 268<br>(67.8%) | 533<br>(72.3%) |  |
| **Diagnostic test results** |  |  |  |  |  |  |  | <0.001 |
| PCR positive | 45<br>(44.6%) | 103<br>(100.0%) | 46<br>(97.9%) | 35<br>(89.7%) | 38 (73.1%) | 355<br>(89.9%) | 622<br>(84.4%) |  |
| IgM positive | 55<br>(54.5%) | 0 (0.0%) | 1 (2.1%) | 4 (10.3%) | 14 (26.9%) | 26 (6.6%) | 100<br>(13.6%) |  |
| Antigen positive | 0 (0.0%) | 0 (0.0%) | 0 (0.0%) | 0 (0.0%) | 0 (0.0%) | 7 (1.8%) | 7 (0.9%) |  |
| **Healthcare challenges** |  |  |  |  |  |  |  | <0.001 |
| Ribavirin stockout | 10 (9.9%) | 17<br>(16.5%) | 0 (0.0%) | 0 (0.0%) | 0 (0.0%) | 92<br>(23.3%) | 119<br>(16.1%) |  |
| PPE shortage | 23<br>(22·8%) | 82<br>(79·6%) | 46<br>(97·9%) | 39<br>(100·0%) | 52<br>(100·0%) | 383<br>(97·0%) | 625<br>(84·8%) |  |

*Note: Data are n (%) unless otherwise specified. IQR=interquartile range. CCHF=Crimean-Congo haemorrhagic* *fever. p values from Kruskal-Wallis test (continuous) or chi-squared test (categorical).*

Binomial proportions are reported with 95% confidence intervals computed using the Wilson score method. A two-sided p-value below 0·05 was considered statistically significant throughout. All models were fitted on complete cases, and the analytic denominator is reported for each. No adjustment was made for multiple comparisons: the primary hypothesis was pre-specified, and the severity-stratified and subgroup analyses are reported as exploratory. Because these comprise a substantial number of comparisons, nominally significant subgroup findings carry an inflated probability of type I error and are interpreted as hypothesis-generating rather than confirmatory throughout.

### Geographic analysis

Patient residence data were mapped across Afghanistan’s 34 provinces using administrative boundary shapefiles from the Global Administrative Data (GADM) database via the geodata package in R. Provinces were grouped into six geographic regions based on Afghanistan’s established administrative structure. Regional hospitalization fatality ratios (HFRs) were defined as the proportion of deaths among all hospitalized laboratory-confirmed patients, were calculated for patients residing in each region across all study hospitals combined (Supplementary Figure 1). Spatial analyses were conducted in R (Version 4.3; R Core Team, 2026) using the sf and ggplot2 packages.

### Reported exposure route analysis

Tick exposure was self-reported by patients or proxies at admission. Occupational categories were pre-specified in the case record form: farmer or livestock handler; healthcare worker (HCW); housewife or household caregiver including meat handling; slaughterhouse worker or butcher; and jobless or other. To describe the reported exposure, we calculated the proportion reporting tick exposure within each occupational group and compared aggregate exposure patterns across hospitals (Supplementary Table 1).

### Healthcare associated infection analysis

Healthcare workers were identified by occupational classification in the case record form. For each healthcare worker case, we defined a likely exposure window as the 5-6 days preceding symptom onset, consistent with the reported CCHF incubation period following contact exposure [1, 5, 7, 24]. We enumerated all non-healthcare worker CCHF patients concurrently hospitalised at the same facility during each healthcare worker’s exposure window. To test whether healthcare worker infections clustered temporally during periods of elevated CCHF patient volume, we performed a permutation test (1,000 iterations) comparing the observed mean number of concurrent CCHF inpatients during healthcare worker exposure windows to a null distribution generated by randomly reassigning healthcare worker symptom-onset dates within the observed range of patient hospitalisations at each facility. A two-sided p-value below 0·05 was considered evidence of non-random temporal clustering.

### Ribavirin-mortality association analysis

Ribavirin receipt was defined using the date of ribavirin initiation recorded in the case record form; patients with a recorded ribavirin start date were classified as treated, including those who received only an initial dose or were treated for one day or less. Ribavirin timing was calculated as days from hospital admission to treatment initiation Treatment duration was not used as an exposure variable because it is truncated by death and is therefore partly a consequence rather than a determinant of outcome. Ribavirin dose was not analysed because dose units were inconsistently recorded across sites.

The primary outcome was in-hospital mortality. We estimated the association between ribavirin receipt and mortality using a multivariable Cox proportional hazards model with time from hospital admission to death or discharge as the survival time. The conventional model adjusted for clinical severity classification (i.e., mild, moderate, severe, further detailed below), age, sex, hospital, and pre-existing comorbidity. Hospital was included as a fixed effect to control for facility-level case-mix confounding identified through exploratory analysis (Supplementary Table 3). The proportional hazards assumption was assessed using Schoenfeld residuals to assess the temporal stability of specific covariate effects.

To investigate discordance between the unadjusted and adjusted mortality estimates, we conducted a stratified log-rank test stratifying by hospital, which provides a non-parametric assessment of the ribavirin survival association after controlling for hospital case-mix without parametric model assumptions (Supplementary Table 4). A logistic regression model was used to examine whether the probability of ribavirin receipt differed by hospital., ruling out reverse causation as an alternative explanation for the adjusted estimate.

Unadjusted Kaplan-Meier survival curves were generated for ribavirin-treated and untreated patients overall and within each clinical severity stratum, with log-rank tests used to assess unadjusted differences in survival. These unadjusted analyses are presented in Supplementary Methods and Supplementary Figures 3-4 to illustrate the extent and source of discordance between crude and adjusted estimates.

### Sensitivity and bias analyses

A large proportion (85%) of patients received ribavirin and treatment was defined by a recorded start date, classification as treated required survival to the point of treatment, creating a risk of immortal-time bias. It was addressed with a hierarchy of pre-specified analyses. The primary analysis compared patients treated on the day of admission (for whom exposure and follow-up both began at time zero, so that no immortal time can arise) with patients who never received ribavirin. Additionally, ribavirin was modelled as a time-varying exposure using the Mantel–Byar approach, splitting each patient’s follow-up at the recorded treatment time so that person-time before treatment was classified as untreated; this analysis was repeated with the time origin set at symptom onset with left truncation at admission. Landmark analyses at days 1, 2, and 3, reclassifying exposure according to treatment status at each landmark, were performed as further sensitivity analyses. Effect modification by the interval from symptom onset to admission was tested to assess a possible therapeutic window. For all models the estimate with and without the treating-hospital term is reported, and baseline characteristics of treated and untreated patients were compared to establish the direction of confounding by indication. The conventional model treating ribavirin as a fixed baseline covariate is reported only to quantify the magnitude of immortal-time bias

### Mortality predictor analysis

To identify independent admission-level predictors of in-hospital mortality, we fitted a multivariable Cox proportional hazards model using only variables available at the time of hospital admission, excluding ribavirin and all treatment variables to avoid reverse causality. Candidate predictors were selected based on clinical plausibility and published CCHF prognostic literature: age, sex, clinical severity category, admission platelet count (per 10,000/μL), Aspartate Aminotransferase (AST) greater than 200 U/L, any haemorrhagic symptom at admission (haematemesis, melaena, epistaxis, haematuria, or petechiae or purpura), neurological involvement (confusion or seizures), and pre-existing comorbidity. White Blood Cell (WBC) count below 3×10⁹/L was identified as a candidate but was present in only three patients and was therefore excluded from the multivariable model due to insufficient power. The model was additionally adjusted for hospital as a fixed effect. Discriminatory performance was assessed using the concordance statistic. To confirm that identified predictors reflected intrinsic disease severity rather than differential treatment access, we assessed hazard ratio stability by comparing estimates with and without ribavirin in the model and repeated the analysis in the ribavirin-treated cohort only.

### Severity classification analysis

To assess whether haemodynamic and laboratory markers improve mortality discrimination beyond the clinical severity classification, we constructed a series of logistic regression models with in-hospital death as the outcome. Models were built incrementally: clinical severity classification alone; plus circulatory failure markers (admission platelet count, vasopressor requirement, blood transfusion, and platelet transfusion); plus hepatic and haematological laboratory markers (AST greater than 200 U/L, WBC below 3×10⁹/L, haemoglobin below 10 g/dL); and plus bedside clinical markers (haemorrhagic symptoms, neurological involvement, oxygen requirement, and comorbidity). Hospital was excluded from all classification models as it is not a patient-level severity criterion. Discriminatory performance was quantified using the area under the receiver operating characteristic (ROC) curve. Differences between models were assessed using DeLong’s test for paired receiver operating characteristic curves. A separate within-stratum analysis was conducted restricted to patients classified as moderate by clinical criteria to assess the extent of severity heterogeneity within this classification.

## Results

### Patient characteristics

A total of 737 patients were hospitalized across the six public hospitals between January 2022 and December 2025. The median age was 35 years (IQR 24–48); 577 (78·3%) were male. Significant variation in patient age, sex distribution, and occupational profile was observed across hospitals (all p < 0·05, Table 1 and Figure 1A). Among 204 CCHF deaths, documented causes (n=195, 95%) predominantly involved overlapping pathophysiologic mechanisms, with sepsis-multiorgan failure being the most common combination (45, 23%), followed by isolated hemorrhagic shock (35, 18%) and the triad of sepsis, hemorrhagic shock, and multiorgan failure (27, 14%) (Figure 1a). The hospitalization fatality ratios (HFRs) which is distinct from the population case fatality rate, and cannot be compared directly with it, differed markedly by hospital, ranging from 8·7% at Herat Infectious Disease Hospital to 40·6% at Balkh Regional Hospital (overall 27·7%; p < 0·001; Table 1). Only 64 study subjects (8·7%) reported having any known tick exposure.

**Figure 1.**
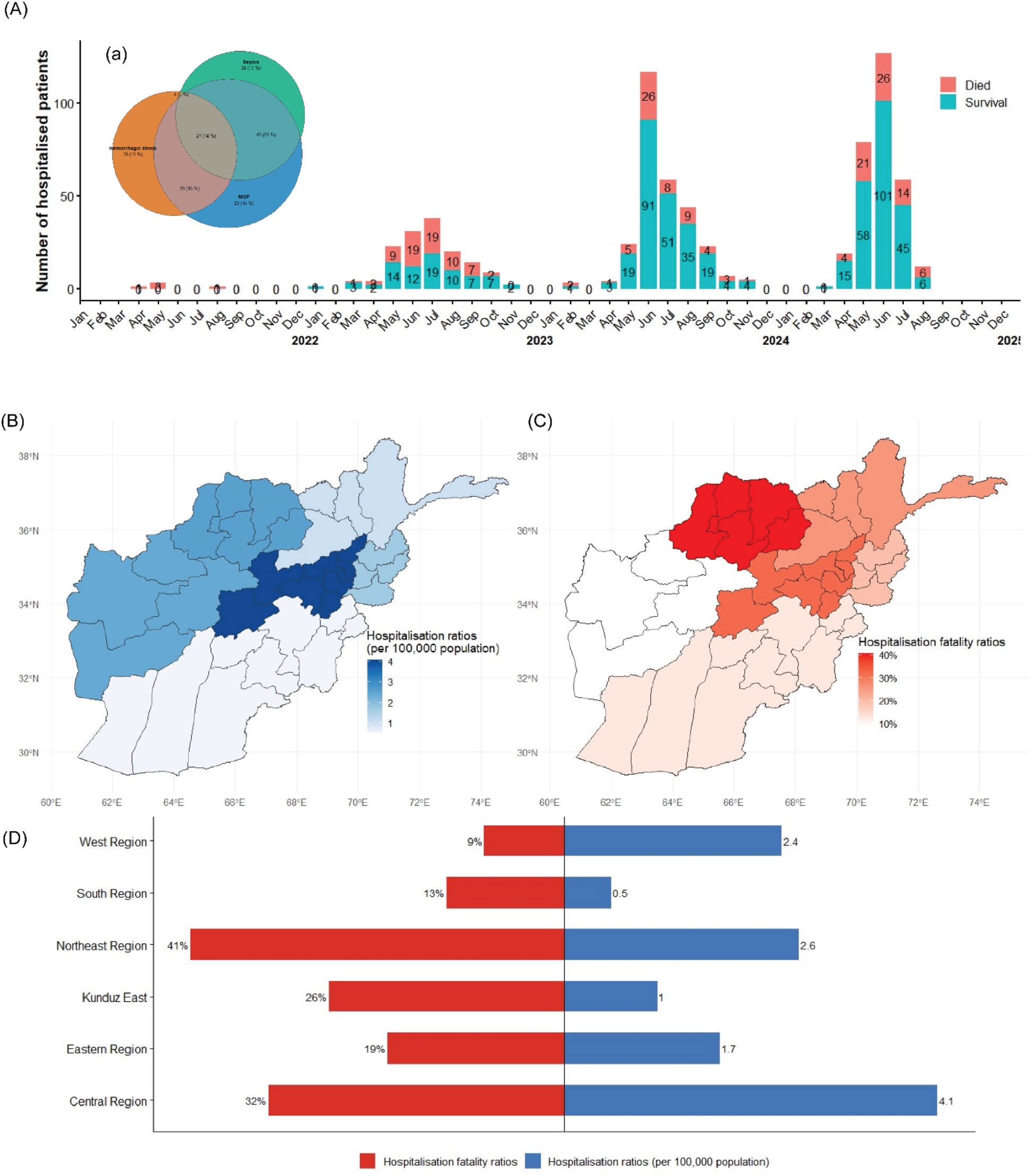
Temporal epidemic curve and mortality outcomes of hospitalised CCHF patients across six Afghan hospitals, 2022–2025. (A) Monthly count of hospitalised patients by outcome (survived, died). (a) Inset: cause-of-death distribution among 195 documented fatal cases. (B-D) Hospitalization ratios per 100,000 population, hospitalization fatality ratios and its corresponding contrasts by region.

All 737 patients underwent RT-PCR, which was positive in 622 (84·4%). Of the 115 RT-PCR-negative patients, 98 were confirmed by IgM serology and 6 by antigen detection alone; a further 6 were IgG-positive only, and 5 had no positive confirmatory result (Table 1). Overall, 100 patients (13·6%) were IgM-positive and 7 (0·9%) antigen-positive, including 3 patients positive on more than one assay. RT-PCR confirmation rates varied substantially by hospital, ranging from 44·6% at Balkh Regional Hospital to 100·0% at Herat Infectious Disease Hospital. Other facilities achieved RT-PCR confirmation rates of 97·9% (Kunduz Regional Hospital), 89·9% (National Infectious Disease Hospital), 89·7% (Kandahar Regional Hospital), and 73·1% (Nangarhar Regional Hospital). IgM serology served as the predominant diagnostic modality at Balkh Regional Hospital (55/101 patients, 54·5%), compensating for the low RT-PCR confirmation rate at this facility. IgM testing also contributed to case confirmation at Nangarhar Regional Hospital (14/52, 26·9%), Kandahar Regional Hospital (4/39, 10·3%), and National Infectious Disease Hospital (26/395, 6·6%). Antigen detection was performed exclusively at the National Infectious Disease Hospital, confirming 7 of 395 cases (1·8%) at this facility (Table 1).

### Seasonal trends of CCHF hospitalization

Marked seasonality was observed across all three epidemic seasons, with peak admissions consistently occurring between May and August (Figure 1A). Admissions increased substantially over the study period: monthly admissions peaked at 3 patients in May 2022, 38 patients in July 2023, 117 patients in June 2024, rising to 127 in June 2025. Among 195 of 204 fatal cases with documented cause of death, sepsis (26·2%), multi-organ failure (20·5%), and haemorrhagic shock (8·8%) were the principal attributed causes, with the majority of deaths involving comorbidities (Figure 1B). HFRs varied substantially by hospital and year (Figure 1C). Balkh Regional Hospital recorded the highest mortality ratio across all seasons (approximately 40%), whereas Herat Infectious Disease Hospital consistently recorded the lowest (approximately 9%).

### Clinical presentations and treatments

Of the symptoms, fever (734/737, 99.6%), headache (713/737, 96.7%), myalgia (669/737, 90.8%), and fatigue (625/737, 84.8%) were the most prevalent, which were jointly present in 566 patients (76.8%), of whom 180 ultimately died (31.8% mortality rate). Haemorrhagic manifestations were common: epistaxis was reported in 230 patients (31.2%), haematemesis in 99 (13.4%), petechiae or purpura in 259 (35.1%), melena in 67 (9.1%), and haematuria in 38 (5.2%). Any hemorrhagic manifestation was present in 499 patients (67·7%) with 176 deaths (35·3% mortality). Neurological features including confusion (337/737, 45·7%) with 131 deaths (38·9% mortality) and seizures (161/737, 21·8%) (Figure 2A).

**Figure 2.**
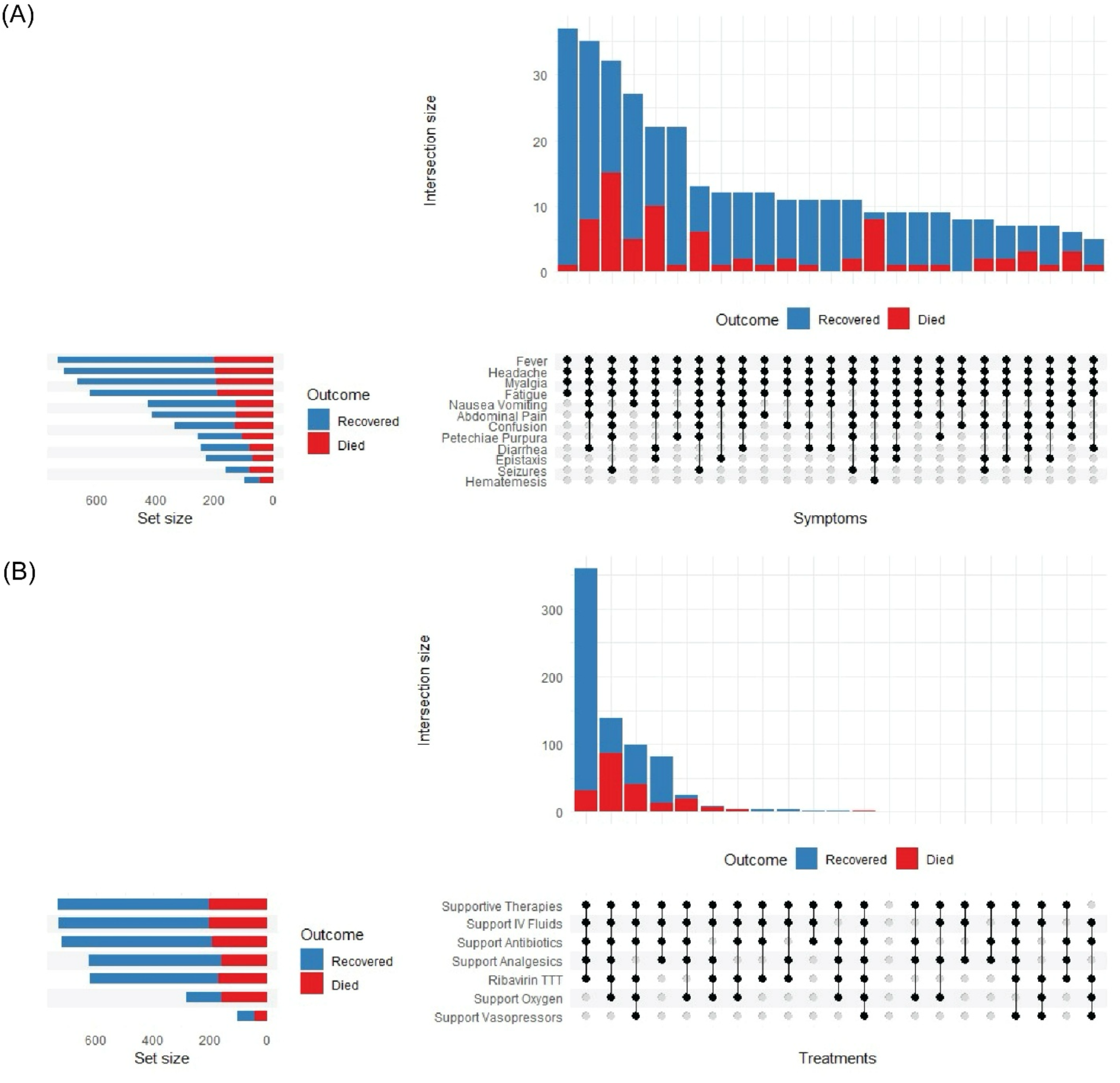
UpSet plots depicting symptom and treatment co-occurrence patterns among hospitalised patients with laboratory-confirmed CCHF, stratified by outcome. (A) Symptom combinations: intersection bars (upper panel) show patient counts for each symptom combination, ordered by frequency. Set size bars (left panel) indicate overall symptom prevalence. Connected dots (matrix panel) identify symptoms comprising each intersection. (B) Treatment combinations: similar layout showing co-occurrence of supportive therapies and ribavirin treatment. Blue bars represent recovered patients; red bars represent deaths.

Symptom intersections involving confusion, seizures, petechiae, and haematemesis were significantly more commonly associated with fatal outcomes (Figure 2A). High-risk of death combinations included confusion plus seizures (80 patients, 49·7% mortality, RR 1·6) and petechiae plus haematemesis (36 patients, 66·7% mortality, RR 2·1) compared to the constitutional tetrad (31·8% mortality). Patients with both hemorrhagic and neurological symptoms had the highest mortality risk (274 patients, 44·9% mortality, RR 3·6) compared to those with constitutional symptoms only (137 patients, 12·4% mortality). Treatment patterns showed that supportive therapies were commonly used in combination, with ribavirin administration frequently accompanied by intravenous fluids, oxygen support, and other supportive measures (Figure 2B).

### Healthcare system challenges

Documented healthcare system challenges were nearly universal and reported amongst almost all hospitalized patients (733/737, 99·5%). Ribavirin stockouts were experienced by 119 patients (16·1%), with the highest burden at the National Infectious Disease Hospital (92/395, 23·3%) and Herat Infectious Disease Hospital (17/103, 16·5%). A documented PPE shortage was recorded for 625 patients (84·8%), reported at every site and universally at Nangarhar (52/52) and Mirwais (39/39). A documented ICU shortage for 399 (54·1%), and cultural barriers to care for 207 (28·1%). The scarcity of organ support was near-absolute rather than relative: among 736 patients with the field recorded, one (0·1%) was admitted to intensive care, and none received mechanical ventilation or dialysis at any site during the four-year period. Supportive care therefore consisted almost entirely of (731/735, 99·5%), antibiotics (722/736, 98·1%), oxygen (282/735, 38·4%), vasopressors (103/736, 14·0%), and blood products (235/734, 32·0%). Access to blood products was the one component of supportive care that differed substantially between sites, ranging from 74·3% of admissions at Balkh to 11·5% at Nangarhar. These systemic deficits likely contributed to the elevated HFRs observed at Balkh Regional Hospital (40·6%) relative to other sites (Table 1). The elevated HFR at Balkh Regional Hospital (40·6%) relative to other sites (Table 1) did not follow the pattern of documented resource deficits, as Balkh had the highest access to blood products and the lowest documented PPE shortage, and may reflect differences in case mix, referral pattern and diagnostic practice.

### Patient-reported risk factors

Although only a minority of patients reported exposure to ticks (64/737, 8·7%), more than half of patients with recorded occupation reported potential occupational or domestic exposures involving regular contact with animal blood or tissues. Occupation was recorded for 574 of 737 patients (77·9%); the following proportions are expressed over that denominator, including: farming or handling of livestock (193/574, 33·6%); slaughterhouse or butchering work (74/574, 12·9%); or other household exposure involving domestic meat preparation (143/574, 24·9%). Tick exposure was most commonly reported by among farmer-livestock handlers (31/193, 16·1%) and shepherds (1/7, 14·3%), followed by slaughterhouse workers (9/74, 12·2%), butchers, and housewives (12/143, 8·4%), who nonetheless comprised a substantial fraction of cases (Supplementary Table 1). Because occupation was missing for 163 patients (22·1%) and tick-exposure context was missing for 681 (92·4%), these distributions describe reported exposures among patients with available data and should not be interpreted as confirmed transmission routes. Spatial variation in reports of tick exposure was substantial. More than half of patients at Kunduz and Kandahar hospitals reported tick bites (53·2% and 51·3%, respectively) compared to none at Balkh and Nangarhar regional hospitals (p < 0·001).

### Healthcare associated infection clustering among healthcare workers

Nineteen of 737 patients (2·6%) were Health Care Workers (HCWs). Infections were geographically concentrated: 13 of 19 (68·4%) occurred at Balkh Regional Hospital, despite that facility treating only 13·7% of the total cohort (101/737; Table 1). Two distinct outbreak periods were observed at Balkh: June 2024 (2 HCW infections during 22 admissions, attack rate 9·1%) and April-July 2025 (11 HCW infections during 47 admissions, attack rate 23·4%). At the National Infectious Disease Hospital, four HCWs were infected during a May–June 2025 surge with 111 admissions over the two-month period (attack rate 3·6%) and up to 49 concurrent CCHF patients. Single cases occurred at Kandahar Regional Hospital (June 2025; 1 HCW infection during 25 admissions, attack rate 4·0%) and Nangarhar Regional Hospital (June 2024; 1 HCW infection during 4 admissions, attack rate 25·0%).

Temporal clustering analysis demonstrated that 18 of 19 HCW infections (94·7%) occurred while at least one CCHF patient was concurrently hospitalized at the same facility (exposure window 3-10 days before HCW symptom onset). The median number of concurrent CCHF patients during HCW exposure windows was 8 (IQR 3-14; range 0-49). The single HCW with no concurrent patient admissions during their exposure window had no reported tick exposure and died. The permutation test indicated that HCW infections clustered significantly in periods of elevated patient volume (observed mean concurrent patients: 11·4 vs. expected: 8·7; p = 0·012), consistent with increased healthcare-associated exposure during outbreak surges; specific transmission events were not documented, and needlestick, splash and mucosal exposures were not recorded (Supplementary Table 2, Supplementary Figure 2).

Personal protective equipment was used in 726 of 736 admissions (98·6%), but its composition was incomplete at most sites: 532 admissions (72·2%) were managed with mask and gloves alone, and only 205 (27·8%) with mask, gloves, and gown. No use of respirators or face shields was recorded at any site. Gown provision was universal at Balkh (101/101) and, Kandahar (39/39), partial at Herat (65/103), and absent at Kunduz, Nangarhar, and the National Infectious Disease Hospital. Isolation rooms were available for 274 of 736 admissions (37·2%), almost all of them at the National Infectious Disease Hospital (259/394, 65·7%); no isolation room was available at Balkh (0/101), Kandahar (0/39), or Nangarhar (0/52). Hand hygiene, waste disposal, disinfection, and injection safety procedures were recorded as in place for between 99·5% and 100% of admissions. The distribution of HCW infection did not follow the distribution of PPE provision. Balkh, where 13 of 19 HCW infections occurred, recorded universal gown use and the lowest documented PPE-shortage rate in the cohort (23/101 admissions, 22·8%), but had no isolation capacity and the highest patient volume-adjusted attack rate. Conversely, the National Infectious Disease Hospital recorded no gown use and near-universal documented PPE shortage, yet four HCW infections across 395 admissions. Fourteen of the 19 infected HCWs were managed at sites where gowns were in use. Taken together, these site-level observations suggest that isolation capacity and caseload may be more closely related to healthcare-associated risk than PPE supply alone, although this was not formally tested and staffing, invasive procedures and sharps handling were not recorded.

### Ribavirin-morality association

Of 737 patients with confirmed CCHF admitted across six hospitals, 10 were excluded due to missing admission or outcome dates (n=8 missing date of outcome, n=2 missing date of admission), and a further 2 because ribavirin status was not recorded, leaving 725 patients in the ribavirin analysis. Of these, 618 (85·2%) received ribavirin and 107 (14·8%) did not. Among 617 treated patients with both admission and treatment start dates recorded, ribavirin was initiated on the day of admission in 442 (71·6%; median day 0, IQR 0–1). The median interval from symptom onset to ribavirin initiation was 5 days (IQR 3–7), mirroring the median onset-to-admission interval of 5 days (IQR 3–7). Treatment duration, reported descriptively, was 10 days or longer in 202 of 618 treated patients (32·7%), 5–9 days in 250 (40·5%), 2–4 days in 106 (17·2%), and 1 day or less in 45 (7·3%), with 15 (2·4%) not recorded. Mortality rose steeply as recorded duration fell (42/45, 93·3% among those treated for ≤1 day versus 25/202, 12·4% for ≥10 days), a gradient that reflects truncation of treatment by death rather than a dose-response relationship. The conventional multivariable Cox proportional hazards model, adjusted for clinical severity, age, sex, hospital, and comorbidity, treating ribavirin as a fixed baseline covariate suggested lower mortality hazard (HR 0·57, 95% CI 0·38–0·85; p=0·006; C-statistic 0·753) (Figure 4), but this estimate is vulnerable to immortal-time bias and is presented for comparability with prior studies. The proportional hazards assumption was satisfied for all covariates and globally (Schoenfeld residual test, global p=0·50, Figure 3). Unadjusted Kaplan-Meier curves showed crossing of survival functions and a non-significant log-rank test overall (p=0·30) and within the Moderate clinical severity stratum (p=0·348), consistent with hospital-level differences in case mix and resources. (Supplementary Figures 3-4).

**Figure 3.**
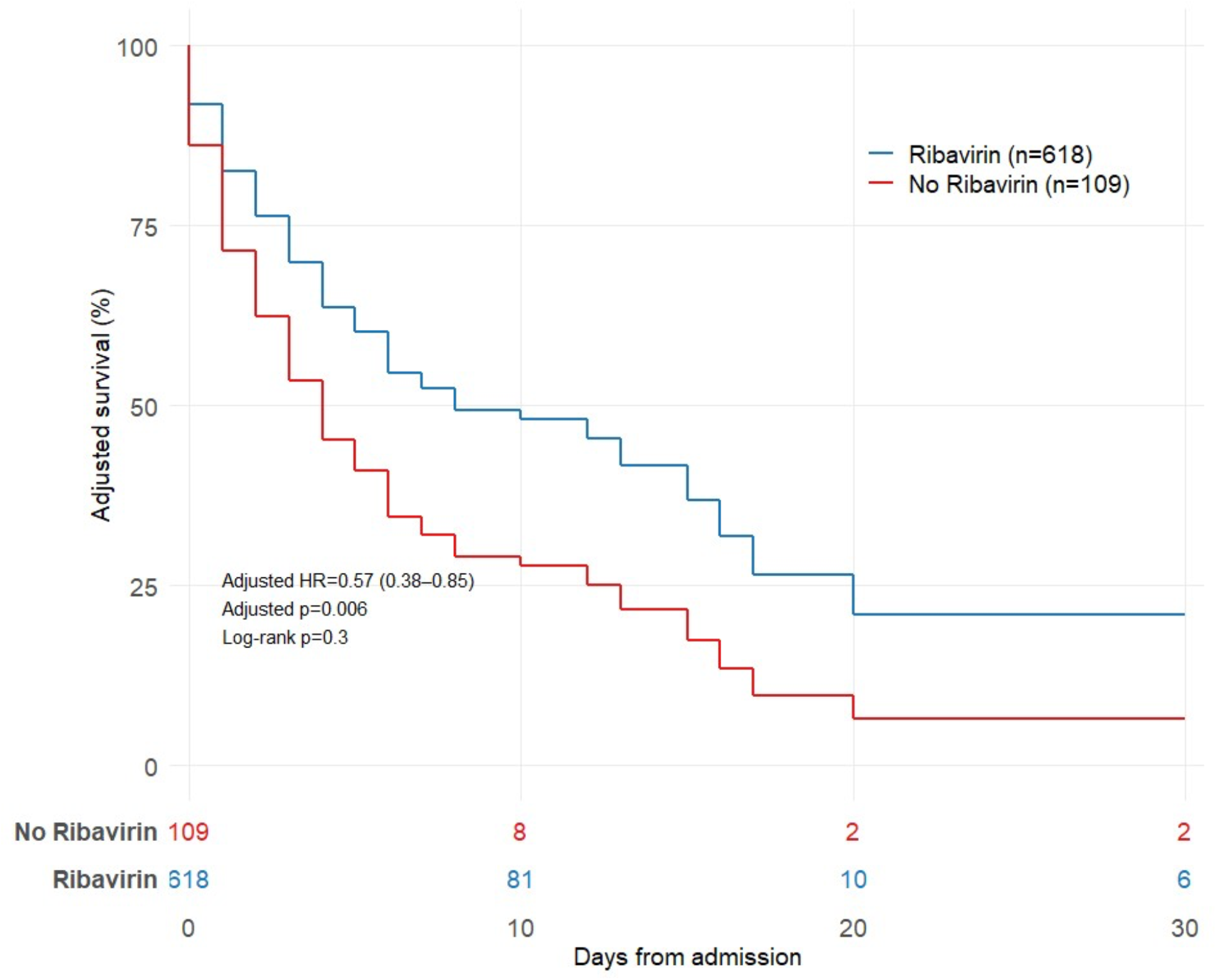
Covariate-adjusted survival curves by ribavirin receipt among patients with confirmed Crimean-Congo haemorrhagic fever (n=727). Survival curves are estimated from the multivariable Cox proportional hazards model adjusting for clinical severity classification, age, sex, hospital, and pre-existing comorbidity, with all covariates fixed at reference values (Mild severity, median age, female sex, most common hospital, no comorbidity). Blue: ribavirin-treated (n=618); red: untreated (n=109). Numbers at risk are shown beneath the x-axis at days 0, 10, 20, and 30. The unadjusted log-rank test was non-significant (p=0·30), reflecting hospital-level case-mix confounding identified at Balkh Regional Hospital (see Supplementary Table 3). In this model, which treats ribavirin as a fixed baseline covariate, ribavirin receipt was associated with an adjusted HR of 0·57 (95% CI 0·38–0·85; p=0·006). As detailed in the main text and Figure 4, this estimate is substantially inflated by immortal-time bias and should not be interpreted as a treatment effect. The proportional hazards assumption was satisfied for all covariates and globally (Schoenfeld residual test, global p=0·50).

**Figure 4.**
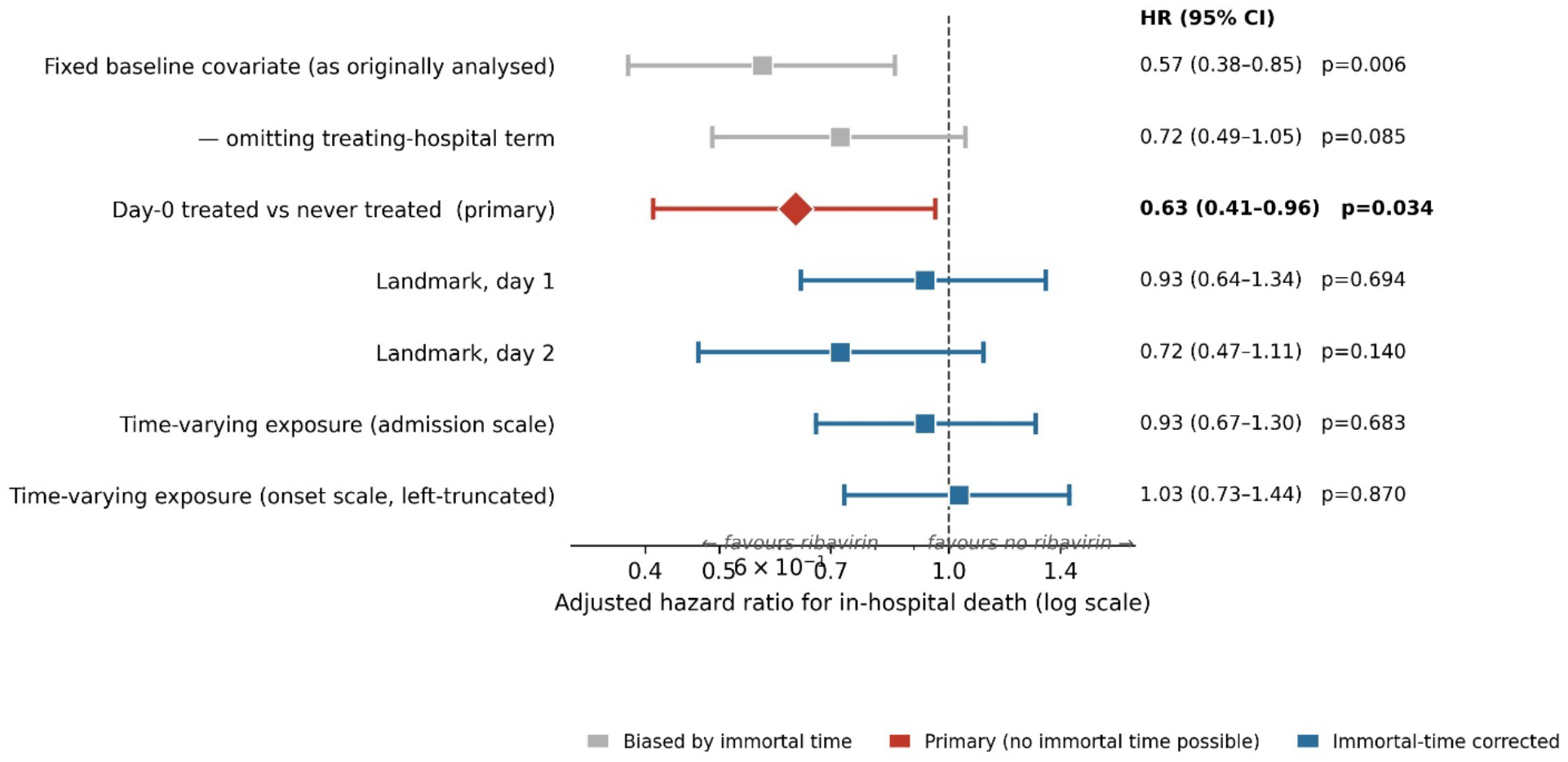
Sensitivity of the estimated ribavirin–mortality association to the handling of treatment exposure. Adjusted hazard ratios (95% CI) for in-hospital death associated with ribavirin are shown for seven pre-specified specifications, all adjusted for clinical severity, age, sex, treating hospital, and comorbidity unless otherwise noted. Grey squares denote analyses in which ribavirin is treated as a fixed baseline covariate and are therefore subject to immortal-time bias, because classification as treated requires survival long enough to receive the drug: the conventional model (HR 0·57, 95% CI 0·38–0·85) and the same model omitting the hospital term (HR 0·72, 95% CI 0·49–1·05). The red diamond denotes the primary analysis, comparing patients treated on the day of admission with those never treated, for whom no immortal time can arise (HR 0·63, 95% CI 0·41–0·96). Blue squares denote immortal-time-corrected analyses: landmark analyses at days 1 and 2 (HR 0·93 and 0·72), and ribavirin modelled as a time-varying exposure on the admission scale (HR 0·93, 95% CI 0·67–1·30) and on the symptom-onset scale with left truncation (HR 1·03, 95% CI 0·73–1·44). The dashed vertical line indicates HR=1·00 (no effect). The apparent benefit under the conventional model is progressively attenuated as immortal time is removed. Severity-stratified Kaplan–Meier curves and the subgroup forest plot previously shown here are provided as Supplementary Figure 4.

The unadjusted log-rank test was non-significant (p=0·30), reflecting hospital-level case-mix confounding. Balkh Regional Hospital contributed 34 patients classified as moderate by clinical criteria who had universal vasopressor requirements (100%), a median admission platelet count of 23,500/μL, and a HFR of 61·8% (21/34) that substantially exceeds that of patients classified as severe at other institutions (39·6%) and is consistent with haemodynamic instability not captured by the haemorrhage-based clinical severity criteria. Ribavirin receipt did not predict hospital allocation within the moderate clinical severity stratum (logistic regression p=0·607), indicating that the between-hospital contrast was explained by differential prescribing within that stratum (Supplementary Table 3-4).

### Severity-stratified ribavirin analyses

These subgroup hazard ratios derive from the conventional baseline-covariate model and share its immortal-time bias; they are presented as exploratory and are not adjusted for multiple comparisons.

Untreated patients were not systematically more severely ill than treated patients at presentation. Among the 111 patients who did not receive ribavirin, none required vasopressors (0% versus 16·5% of treated patients), median admission platelet count was higher (35,000 versus 26,500/μL), median AST was lower (113 versus 166 U/L), and a smaller proportion were classified as severe (38·7% versus 44·1%). Haemorrhagic manifestations (65·8% versus 68·1%), neurological involvement (45·9% versus 45·7%), and comorbidity (6·3% versus 5·0%) were comparable. Confounding by indication in this cohort therefore operates in the direction of biasing the ribavirin estimate towards the null rather than away from it (Table 2).

The conventional estimate was substantially attributable to immortal-time bias. Among treated patients, ribavirin was recorded as starting after the day of admission in 175 (28%), yet the median length of stay among patients who died was 2 days (IQR 1–4), with 82 deaths within 24 hours of admission; classifying ribavirin at baseline therefore credited the treated group with survival time that necessarily preceded treatment. When ribavirin was modelled as a time-varying exposure, the association was fully attenuated (HR 0·93, 95% CI 0·67–1·30), and this was unchanged when the time origin was set at symptom onset with left truncation (HR 1·03, 95% CI 0·73–1·44). Landmark analyses reclassifying exposure at each landmark gave HR 0·93 (day 1) and 0·72 (day 2), and no effect modification by the onset-to-admission interval was detected (interaction HR 1·05, 95% CI 0·95–1·17), providing no evidence of a therapeutic window. In the primary analysis, comparing patients treated on the day of admission (n=439, for whom no immortal time can arise) with those never treated (n=109), ribavirin was associated with lower mortality hazard (adjusted HR 0·63, 95% CI 0·41–0·96). This estimate was borderline and sensitive to model specification, moving above conventional significance under minor changes; the point estimate was stable at approximately 0·63–0·67 across specifications while its significance was not. The estimate was, as expected, dependent on adjustment for treating hospital, since ribavirin availability and baseline mortality both cluster by site. Taken together, point estimates ranged from 0·57 in the immortal-time-affected fixed model to 0·93–1·03 in the time-varying analyses, and that with 34 deaths among untreated patients the cohort is powered to detect only large effects (HR ≤0·5); the findings are therefore consistent with, but cannot confirm, a clinically meaningful benefit.

### Risk factors at admission of CCHF mortality

In multivariable prognostic analysis of admission characteristics (n=641, events=188, C-statistic 0·807), independent predictors of in-hospital mortality were AST greater than 200 U/L (HR 4·13, 95% CI 2·87-5.93; p<0·001), neurological involvement at admission (HR 2·11, 95% CI 1·43-3·11; p<0·001), any haemorrhagic symptom (HR 2·00, 95% CI 1·17-3·42; p=0·011), and older age (HR 1·01 per year; p=0·023). Leukopenia was present in only three patients and was excluded from the multivariable model. Pre-existing comorbidity and platelet count were not independently associated with mortality in the complete-case dataset. Clinical severity classification did not independently predict mortality after adjustment for these markers (Moderate vs Mild HR 1·28, p=0·483; Severe vs Mild HR 1·89, p=0·079). The three major predictors of mortality (i.e., evidence of hepatic neurological, and haemorrhagic complications) remained significant among ribavirin-treated patients only (n=552; AST HR 4·55, haemorrhage HR 2·13, neurological HR 1·82; all p<0·02), suggesting these markers reflect disease severity rather than treatment access (Supplementary Table 5 and Supplementary Figure 6).

## Discussion

### Exposure and the One Health interface

Tick exposure was reported by only 8·7% of patients, against 51–72% in Turkish clinical series [30–32] and 26–70% in smaller Iranian series [33–34]. This difference can be interpreted as recalled exposure rather than proof of a different transmission ecology: Hyalomma nymphs detach unnoticed, and severely ill or fatally affected patients often cannot provide an exposure history. What the data show positively is that occupational and household animal contact dominated the reported exposures — livestock handling, slaughter, and domestic meat preparation together accounting for most patients with an occupation recorded. Because infected livestock remain asymptomatic, these enzootic cycles go undetected, and interventions at the animal–human interface should therefore be strengthened alongside, not instead of, tick-bite prevention. The rise in monthly admissions over the study period, peaking each year in the one to two weeks after Eid-al-Adha, is consistent with intensified livestock contact during animal sacrifice and meat handling, but this interpretation remains ecological and should be tested prospectively [35–36].

### Nosocomial risk and the health-system context

The permutation analysis provides formal statistical evidence that healthcare-worker infections clustered with patient volume during surges (p=0·012), extending descriptive reports from Iran, Türkiye, and Pakistan [24–25, 37–38]. The distribution of infections suggests that factors beyond PPE supply were relevant. Balkh Regional Hospital contributed 13 of 19 healthcare-worker cases despite treating only 13·7% of the cohort, and recorded the highest HFR (40·6%), the highest attack rate during the April–July 2025 surge (23·4%), and no isolation capacity (0 of 101 admissions). However, because specific exposure events were not documented, these findings establish temporal clustering consistent with healthcare-associated exposure, not a proven transmission route. Possible mechanisms include patient-to-HCW exposure during care, contaminated sharps, blood products, shared equipment and laboratory exposure, which could not be distinguished in these data. PPE composition remains relevant: across the cohort, protective equipment was usually recorded but often incomplete, with mask and gloves without a gown in 72·2% of admissions and no recorded respirator or face-shield use at any site. Completing PPE composition is low-cost and immediately actionable, whereas isolation capacity requires capital investment and lead time; both are needed and neither substitutes for the other.

The context in which this care was delivered warrants emphasis, because it bears directly on interpretation. Among 737 patients with HFR of 27·7%, one patient was admitted to intensive care and none received mechanical ventilation or dialysis at any site over four years. Organ support was, in effect, unavailable. This shapes what the mortality data can and cannot say: differences in intensive care between “better-resourced” and other centres cannot confound our comparisons, because no such gradient existed, and the fatality ratio reported here describes CCHF managed with supportive care alone.

### Ribavirin

Our unadjusted-for-immortal-time estimate (HR 0·57) closely mirrors the pooled relative risk of 0·56 (95% CI 0·35–0·90) from a meta-analysis of observational studies, which was graded as very low certainty because of confounding [26]. The only randomised trial, by contrast, found no mortality benefit (RR 1·13, 95% CI 0·29–4·32), although with 136 participants it was too small to exclude a clinically important effect [39], and a second meta-analysis likewise found no survival advantage (RR 1·06, 95% CI 0·97–1·16) [27]. That a conventional analysis of our cohort reproduces the observational literature so closely suggests that much of the apparent benefit reported previously may reflect the same biases.

We do not interpret these findings as evidence that ribavirin is ineffective. The point estimate in the least-biased analysis remained below one, its significance was borderline and specification-dependent, and with only 34 deaths among untreated patients the study was powered to detect only large effects. The appropriate conclusion is an absence of evidence of large benefit, not evidence of absence; residual confounding by indication — which here would bias towards the null, since untreated patients were marginally less severe — cannot be excluded. Adequately powered randomised trials remain necessary, and observational analyses should model antiviral exposure as time-varying before informing ribavirin allocation policy.

### Severity grading

The clinical severity classification discriminated mortality only moderately (AUC 0·65). Admission AST above 200 U/L was independently associated with death (adjusted HR 4·13, 95% CI 2·87–5·93), consistent with Turkish cohorts in which markedly raised AST predicted death [40], but admission platelet count was not, and we do not propose it as an additional criterion. We had considered vasopressor requirement as a candidate severity marker but withdraw that proposal: vasopressor use was recorded in 99·0% of admissions at Balkh and in 0–2% elsewhere, indicating that this variable reflects site-level documentation practice rather than a patient-level severity gradient and cannot be separated from treating hospital in these data. Prospective, protocol-standardised validation is required before any revision of CCHF severity grading.

### Limitations

This retrospective analysis of routinely collected records supports only observational associations. Several limitations bear on interpretation.

First, the ribavirin comparison rests on a small untreated group (34 deaths), is powered only for large effects, and is subject to residual confounding by indication and to the immortal-time bias detailed above; we therefore report a hierarchy of specifications rather than a single casual estimate. Second, exposure classification is subject to substantial misclassification: tick exposure was self-reported and frequently unrecalled, occupation was missing for 163 patients (22·1%) and tick-exposure context for 681 (92·4%), Reported livestock, slaughtering, butchering, household meat preparation, and occupational exposures should be interpreted as reported histories rather than confirmed routes, and the data cannot distinguish tick exposure, animal exposure, shared household exposure, or secondary household transmission. Third, the healthcare-worker analysis establishes temporal clustering, not a mechanism — needlestick, sharps, splash, and mucosal exposures were not recorded, no serological screening of asymptomatic staff was undertaken, and the biosafety level of sample processing was not documented, so the route of transmission remains unexplained and the attack rate is likely underestimated. Fourth, severity was assigned by attending physicians without central adjudication or a validated instrument, and marked between-site heterogeneity indicates inconsistent thresholds. Fifth, hospital resource differences may have confounded treatment allocation, mortality, and healthcare-worker risk, including differences in isolation capacity, blood products, diagnostic methods, staffing, laboratory capacity, and ribavirin availability. Sixth, all models were complete case, missingness was non-uniform and plausibly related to outcome, and ribavirin dose could not be reported because units were inconsistent across sites. Reasons for non-treatment and for treatment discontinuation were not available, and diagnostic methods differed between sites (IgM serology alone confirmed 54·5% of cases at Balkh). Seventh, this hospitalised tertiary-care cohort excludes pre-hospital and community-managed cases, so the 27·7% fatality ratio reflects ascertainment, referral pattern, case-mix and health-system constraints and should not be compared directly with population case-fatality rates of 5–10% from settings with broader surveillance. Generalizability to non-tertiary settings and to other endemic countries with different referral systems and health-system capacity is therefore limited. Finally, no sample-size calculation was performed, as this was a census of eligible cases; confidence intervals are reported throughout so that precision can be judged directly, and subgroup analyses, unadjusted for multiple comparisons, should be regarded as hypothesis-generating.

### Conclusion

Three priorities follow. First, CCHF prevention in Afghanistan should strengthen animal-human interface interventions while maintaining tick-bite prevention, because reported tick exposure was likely under-ascertained. Practical measures include protective equipment for slaughter workers and butchers, community education for household meat handlers, veterinary surveillance, and pre-Eid-al-Adha risk reduction under a One Health framework spanning the Ministries of Public Health and Agriculture and Environment. Second, healthcare-associated prevention requires investment in dedicated isolation capacity, adequate staffing, complete PPE, safe sharps and blood-exposure practices, and surge protocols at high-volume regional hospitals; PPE supply alone is insufficient. Third, the apparent survival association of ribavirin in observational data should be interpreted cautiously: these findings do not establish efficacy, and treatment policy should rest on adequately powered trials and analyses that account for treatment timing. Afghanistan’s accelerating epidemic and unregulated cross-border livestock movement underscore the need for regional coordination of surveillance, laboratory capacity, isolation capacity, and uninterrupted drug supply.

## Data Availability

All data produced in the present study are available upon reasonable request to the authors

## Declarations

### Contributors

RZ and MNS developed the study protocol and questionnaire. RZ coordinated data collection from the hospitals and the Ministry of Public Health. SC led the data analysis and manuscript development. RZ and MNS contributed to the manuscript. FA and SC edited the first draft. YC, HS, GS, TE, LJW, and NH reviewed the first draft and revised version of the manuscript and provided their inputs. MNS corresponded with all authors throughout the process. All authors reviewed the final draft and approved the final version.

### Data sharing

De-identified individual patient data and analysis code are available from the corresponding author upon reasonable request, subject to local ethics committee approval.

## Acknowledgements

We acknowledge the Ministry of Public Health and the six hospitals for sharing the data and TDR small grant for completion of the study.

